# Harnessing testing strategies and public health measures to avert COVID-19 outbreaks during ocean cruises

**DOI:** 10.1101/2021.01.24.21250408

**Authors:** Gerardo Chowell, Sushma Dahal, Raquel Bono, Kenji Mizumoto

## Abstract

To ensure the safe operation of schools, workplaces, nursing homes, and other businesses during COVID-19 pandemic there is an urgent need to develop cost-effective public health strategies. Here we focus on the cruise industry which was hit early by the COVID-19 pandemic, with more than 40 cruise ships reporting COVID-19 infections. We apply mathematical modeling to assess the impact of testing strategies together with social distancing protocols on the spread of the novel coronavirus during ocean cruises using an individual-level stochastic model of the transmission dynamics of COVID-19. We model the contact network, the potential importation of cases arising during shore excursions, the temporal course of infectivity at the individual level, the effects of social distancing strategies, different testing scenarios characterized by the test’s sensitivity profile, and the testing frequency. Our findings indicate that PCR testing at embarkation and daily testing of all individuals aboard, together with increased social distancing and other public health measures, should allow for rapid detection and isolation of COVID-19 infections and dramatically reducing the probability of onboard COVID-19 community spread. In contrast, relying only on PCR testing at embarkation would not be sufficient to avert outbreaks, even when implementing substantial levels of social distancing measures.

## Introduction

Since the first human infections of the novel coronavirus (SARS-CoV-2) were reported in Wuhan, China in December 2019, the novel pathogen has reached every corner of the world and continues its unrelentless global march with more than 95.6 million reported cases including over 2 million related deaths by January 19^th^, 2021 ^1^. The novel coronavirus has spread with significant geographical distinction at different spatial scales around the world. In the United States, a total of 24.1 million COVID-19 cases including more than 400,000 deaths have been reported thus far ^1^. Although safe and effective vaccines are becoming a reality in an unprecedented time scale, it will take months before a substantial fraction of the world population is immunized.

SARS-CoV-2 is a highly transmissible and deadly respiratory virus that readily spreads via droplets and aerosols especially in confined settings ^2,3^. In fact, an early hotspot of the novel coronavirus outside mainland China unfolded aboard the Diamond Princess Cruise ship with 2,666 passengers and 1,045 crew members. This unfortunate COVID-19 outbreak shed early light on the clinical and epidemiological features of this novel coronavirus ^4-6^. In particular, the outbreak on the Diamond Princess Cruise ship highlighted a substantial frequency of asymptomatic infections which need to be rapidly isolated in order to halt transmission chains ^4^.

Because the COVID-19 pandemic has greatly disrupted economic growth, there is an urgent need to develop cost-effective public health strategies that allow safe operation of schools, workplaces, nursing homes, and other businesses ^7-9^. In this paper our focus is the cruise industry which was hit early by the COVID-19 pandemic, with more than 40 cruise ships reporting COVID-19 infections ^10,11^. This form of leisure travel has undergone rapid growth in recent years including a substantial increase in ship size and passenger capacity ^12^. According to the Cruise Line International Association report, the total number of cruise passengers increased from 17.8 million in 2009 to 26.7 million in 2017 and was projected to increase to 30 million in 2019 ^13^.

Multiple infectious disease outbreaks have been linked to cruise ships ^14,15^. A cruise ship mimics a virtual travelling city bringing together a large number of people from different backgrounds, culture, and health status ^16,17^. The interaction of passengers and crew in close proximity in often crowded, semi-enclosed environments such as dining halls and recreational rooms create a unique environment that facilitates the transmission of person-to-person, food borne or water borne diseases ^14^. Some earlier outbreaks of respiratory illness involving cruise ships include an influenza outbreak that occurred in May 2009 where 3% of passengers and crew were infected with A/H1N1 influenza, 3.6% with A/H3N2, and 0.1% with both strains ^18^. Another influenza B outbreak was documented on a cruise ship off the Sao Paulo coast in Brazil in February 2012 ^19^. Similarly, outbreaks of acute respiratory illness were reported in two cruise ships affecting 3.7% and 6.2% of the passengers, respectively, between March 15 and April 5 in 2014 ^20^.

There is a need to devise public health strategies to ensure safe transportation of passengers and crew across different transportation modalities and geographic distances. Indeed, international travel through cruise ships can have significant impact on the transmission and global spread of infectious disease. In the absence of appropriate screening and control measures, infectious individuals who disembark from ships and use multiple transportation means including trains, buses, and international flights can in turn transmit the disease to other people. Additionally, the average passenger on a cruise ship tends to be older and is at heightened risk of severe symptoms and complications from COVID-19 infection ^21^. According to a prospective study, the average age of passengers who sought medical attention in 86 cruises of a ship in three years, was 72.6 years ^22^. In the Diamond Princess Cruise ship, out of total 3,711 people aboard on 5^th^ February 2020, 58.5% were aged 60 years and above, with 33.4% of the individuals aged 70-99 years ^23^. Therefore, ensuring the safety of passengers and the crew aboard cruise ships as well as the local communities that host them is the highest priority of the cruise industry before operations are restarted ^24,25^. To that end, some cruise companies have started to install PCR laboratories aboard their ships with capacity for daily testing of every crew member and guest ^26^.

In this study, we apply mathematical modeling to assess the impact of testing strategies together with social distancing protocols on the spread of the novel coronavirus during ocean cruises using an individual-level stochastic model of the transmission dynamics of COVID-19. We model the contact network of the population of interest and the potential importation of cases arising during shore excursions, our understanding of the temporal course of infectivity at the individual level as well as the effects of social distancing strategies and different testing scenarios characterized by the sensitivity profile of the test and testing frequency.

Our modeling results indicate that PCR testing at embarkation and daily testing of all individuals aboard, together with increased social distancing and other public health measures, should allow for rapid detection and isolation of COVID-19 infections before they infect others, significantly reducing the onboard COVID-19 community spread. Our results support a daily PCR testing strategy in order to minimize the number of infections irrespective of the duration of the cruise, allowing cruises longer than 7 days. By contrast, a strategy that relies on PCR testing at embarkation would not be sufficient to avert outbreaks, even when substantial levels of social distancing measures are implemented.

## Methods

### Model description

#### Model description

We developed an individual-level stochastic model to investigate the role of testing and social distancing protocols for preventing COVID-19 outbreaks during ocean cruises that include daily shore excursions. For this purpose, we model transmission dynamics in a highly connected social contact network and calibrate the baseline transmission rate based on estimates derived from the COVID-19 outbreak that unfolded aboard the Diamond Princess Cruise ship in February 2020 ^4^. Further, our model incorporates uncertainty in the individual-level infectivity profile, which is informed by published data, the role of social distancing measures for mitigating the transmission probability per contact, potential exposure during daily shore excursions based on a local community prevalence, and the uncertainty associated with the sensitivity profile of the PCR test utilized in different testing scenarios.

Through global sensitivity and uncertainty analysis and using 200 stochastic simulations for each combination of parameter values, we summarized our findings in terms of the cumulative number of cases occurring during the duration of the cruise for scenarios with and without the implementation of interventions. A detailed description of the different model components is provided below.

##### Transmission Dynamics and Infectivity Profile

In the absence of interventions, susceptible individuals in contact with infectious individuals become infected with a probability that depends on an infectivity level of each infectious contact that varies according to the age of infection and a scaling factor that modulates transmission rate according to the basic reproduction number R_0_. The infectivity of infectious individuals varies according to their age of infection which progresses on daily time steps (Figure 1). Thus, for a 14-day infectious period, there are 14 different infectivity values for each infectious individual. All infectious individuals recover and become protected at the end of their infectious period.

**Figure 1.**
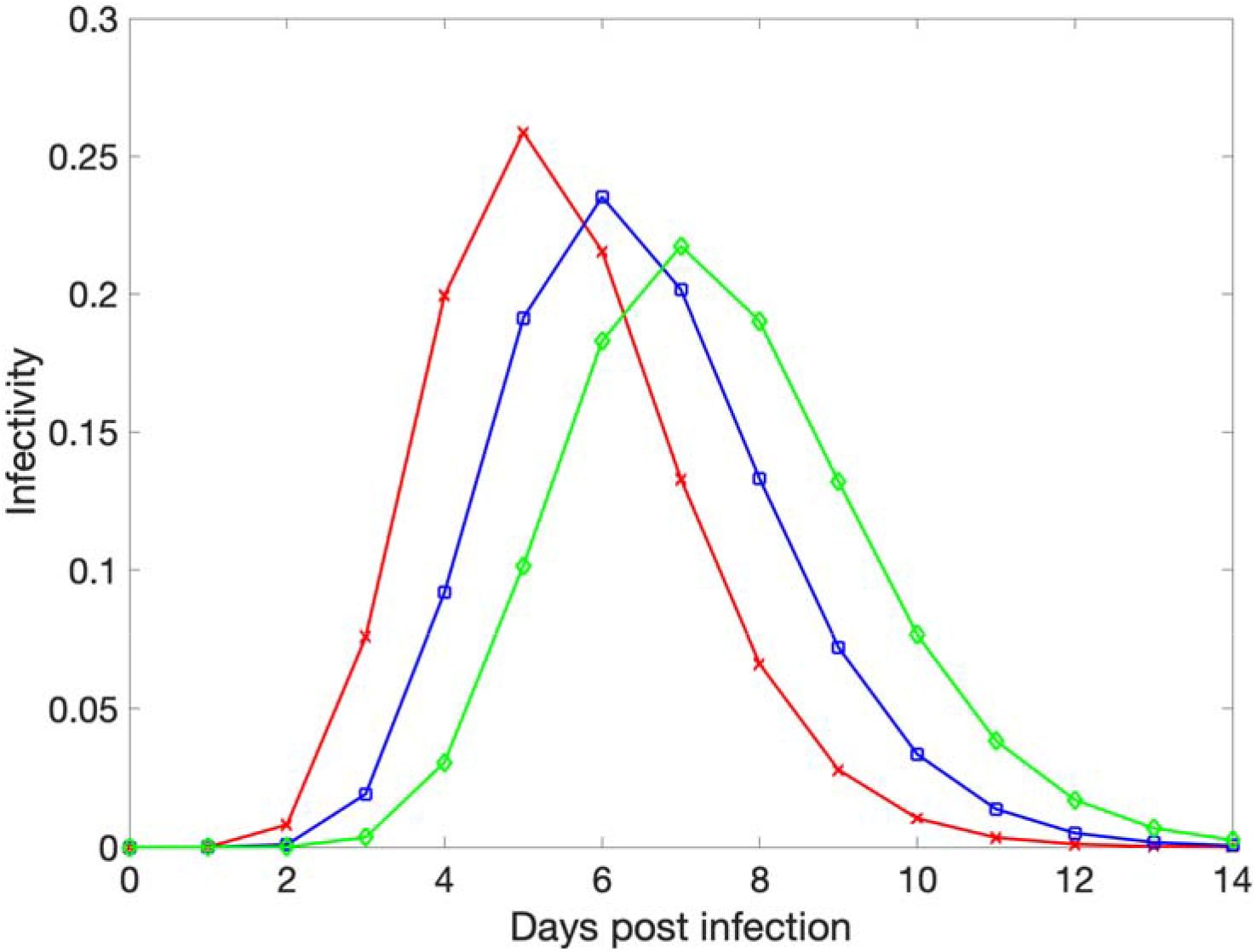
The timing of peak infectivity for each individual varies randomly, peaking at 5 (red curve),6 (blue curve), or 7 (green curve) days post infection according to a gamma distribution. We do not distinguish between symptomatic and asymptomatic individuals.

Asymptomatic transmission is a key characteristic of the transmission dynamics of COVID-19 outbreaks ^27^. Therefore, it is crucial to rapidly identify infected individuals irrespective of the presence of symptoms. While we do not distinguish between symptomatic and asymptomatic individuals in our model, we track and model the uncertainty in the temporal infectiousness profile for each infected individual. For this purpose, we approximate the COVID-19 infectivity profile following a gamma distribution function informed by published data ^28,29^. Specifically, we assume that the infectious period is up to 14 days long, but the timing of peak infectivity for each individual varies randomly and occurs 5, 6, or 7 days post infection as shown in Figure 1.

##### Individual contact network

We model a highly connected network of contacts among passengers and crew aboard the vessel to capture a well-mixed population in a confined setting. For this purpose, the baseline contact network of 1395 individuals including passengers and crew is conservatively modeled according to a small world network where each individual has an average of 100 contacts during the duration of the cruise itinerary (i.e., small world network parameter K=50) with a rewiring probability p set at 0.1 ^30^. The resulting small world network has a low average path length and significant clustering compared to its random network counterpart. Further, the links of the network do not change during the entire duration of the simulation. The baseline contact network is depicted in Figure 2. In sensitivity analyses, we also consider a bigger ship with capacity of 6000 (4000 passengers and 2000 crew).

**Figure 2.**
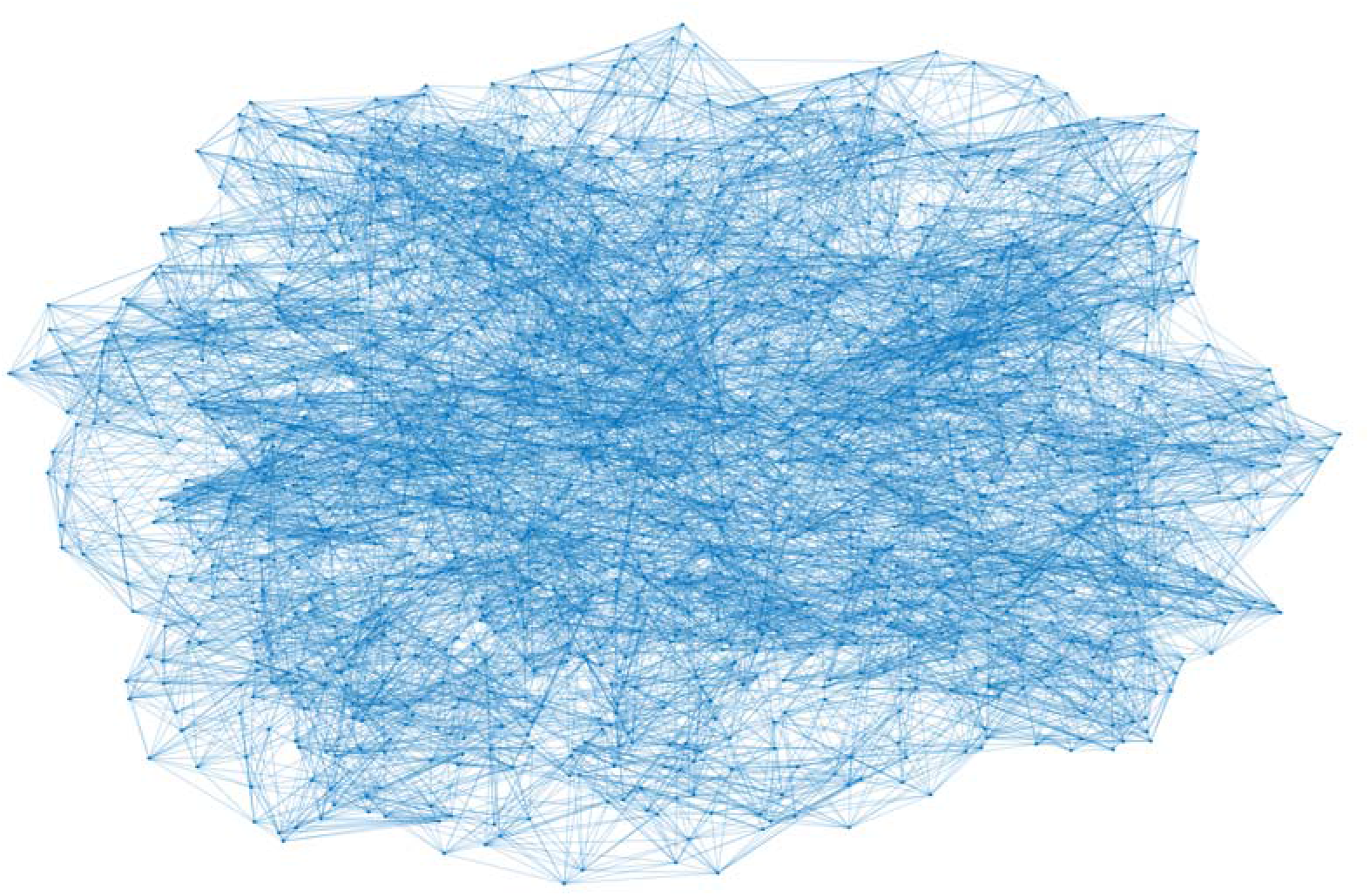
Contact network involving 1395 individuals on board the cruise ship following the small world network model with baseline values (K=8 and p=0.1). The dots represent individuals, and the lines represent fixed contacts between individuals.

##### The basic reproduction number on board the vessel (R_0(ship)_)

The basic reproduction number denoted by R_0_ quantifies the average number of secondary cases generated by a primary infectious during his/her infectious period in the absence of interventions during the early stages of an outbreak ^31^. In line with published studies of the early transmission dynamics of the COVID-19 outbreak that unfolded aboard the Diamond Princess Cruise ship in early 2020 ^5,32^, we calibrated our transmission model with a baseline average basic reproduction number aboard the cruise ship denoted by R_0(ship)_ at 12, but vary this value in the range between 9 and 16 in sensitivity analyses.

##### COVID-19 Test characteristics

We model testing strategies based on PCR tests with a sensitivity profile that is modeled using a logistic growth function that reaches maximum sensitivity 5-7 days post infection followed by a symmetric logistic decline function. Further, we vary peak test sensitivity from 85% to 95% in sensitivity analyses (Figure 3).

**Figure 3.**
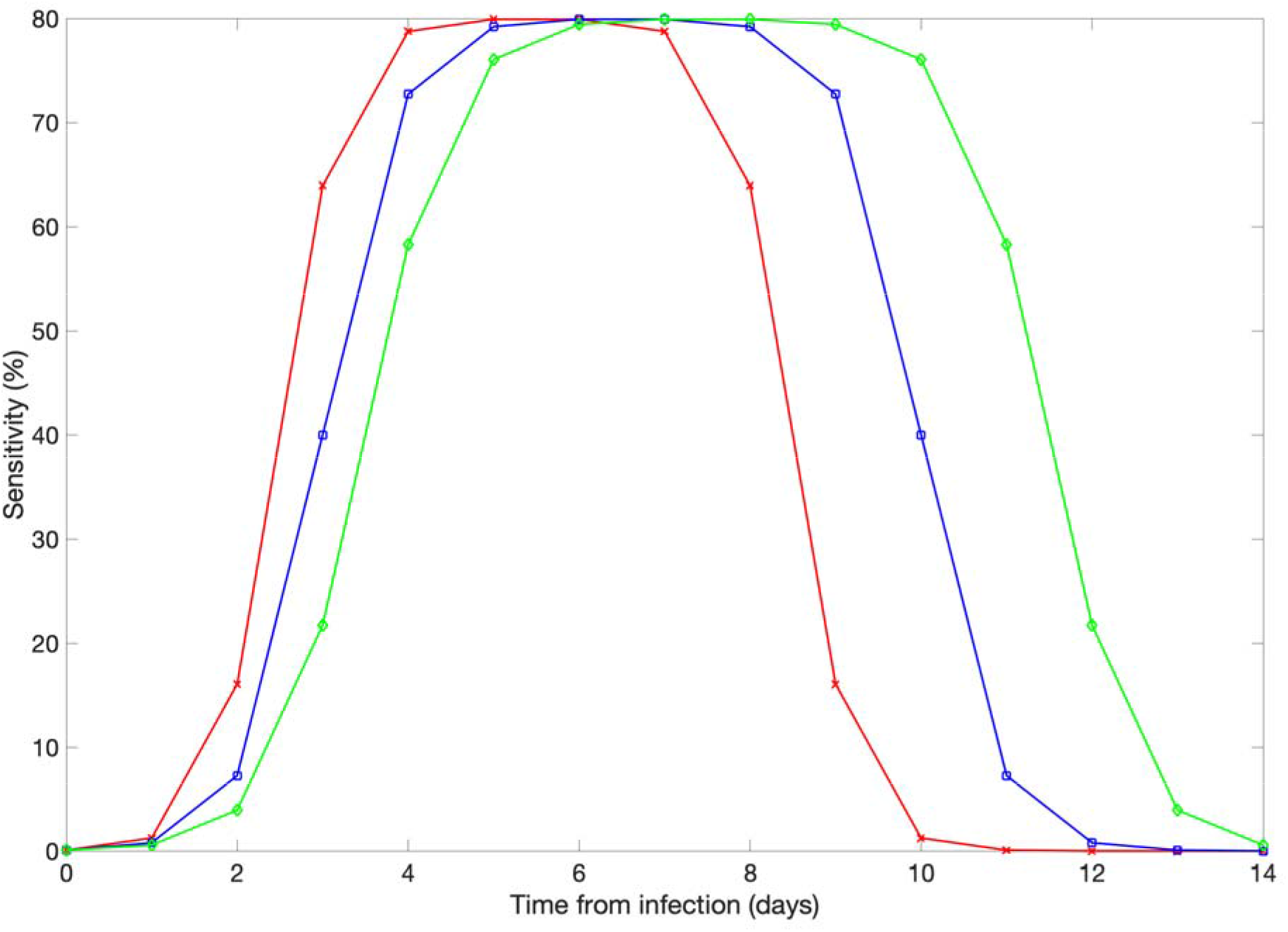
Illustration of the PCR test sensitivity profiles modeled according to a logistic function where a peak sensitivity of 80% is reached 5 (red curve), 6 (blue curve), or 7 (green curve) day post infection.

##### Social distancing protocols

The overall effect of public health measures during the duration of the cruises (including during shore excursions) is modeled as a proportionate reduction in the transmission rate by a scaling factor. These public health measures include frequent sanitation of surfaces, mandatory wearing of mask on board for passengers and crew, physical distancing protocols, air purifiers, and UV lights in all air handling units ^33-36^. Hence, the combined effect of these public health measures are assumed to reduce the transmission probability per contact in the range of 50-90% with a conservative baseline level at 60%.

##### Testing strategies

In addition to social distancing measures, we evaluate the impact of two different testing protocols on the cumulative number of cases that occur during the cruise duration:

- ***PCR Testing of passengers prior to embarkation***. All of the individuals are tested prior to embarkation, and positive individuals are not allowed to board the ship. The probability of a positive individual boarding the ship depends on the age of his/her infection as well as the temporal sensitivity profile of the test. Hence, the probability of a false-negative test result is not negligible ^37,38^. Indeed, even PCR tests are unlikely to detect very recent infections. Infected individuals that are detected prior to embarkation do not contribute to the transmission dynamics during the cruise itinerary.
- ***PCR Testing of passengers prior to embarkation and daily testing on board***. In addition to testing all passengers prior to embarkation, all individuals on board the ship are tested every day with a PCR test. We assume that test results become available within hours and positive cases are effectively isolated and do not contribute to generating further infections on board the ship.

##### Prior immunity

For simplicity we assume that all of the individuals on board the cruise are equally susceptible to catching the novel coronavirus. Thus, we model here conservative scenarios since the potential role of prior immunity from past infection or vaccination campaigns is not considered in our analyses.

##### Initial conditions

The initial number of infected passengers before the cruise starts depends on the local community prevalence with a baseline value at 1% which we vary in the range: 0.3-3% in sensitivity analyses. Specifically, the initial number of COVID-19 infected passengers is drawn from a binomial probability distribution based on the community prevalence value and the number of passengers boarding the ship. Moreover, all crew members are initially assumed to be susceptible to infection with SARS-CoV-2. Moreover, the age of the infection of initially infected individuals at embarkation varies randomly between 0 and 14 days.

##### Daily importation rate from shore excursions

We also consider the possibility that passengers may be exposed to SARS-CoV-2 during daily excursions on shore. For each day of the cruise duration, the number of infected passengers during the shore expedition depends on the current number of susceptible passengers, the local reproduction number on shore denoted by R_0(shore)_ which is assumed to vary between 0.6 and 1.6, the local COVID-19 prevalence level, the amount of time that passengers spend on shore excursions (about 6 hours) as well as the extent of public health measures in place during these activities (e.g., facemask wearing, hand hygiene). Hence, the number of passengers infected during shore excursions could fluctuate on a daily basis depending on the epidemiological state of the individual passengers. Here we assume the same local prevalence during embarkation and during daily shore excursions (range: 0.3-3%). Figure 4 illustrates the average daily number of infected individuals from shore excursions as a function of the local prevalence and the extent of public health measures which we quantify as a proportionate reduction between 50% and 90% in the transmission rate. This is likely a conservative range that reflects the combined impact of mandatory facemask wearing for passengers, physical distancing protocols during the excursion, buses sanitation, limiting of high-risk shore excursion programs and of self-exploration of the destination ^34,36^.

**Figure 4.**
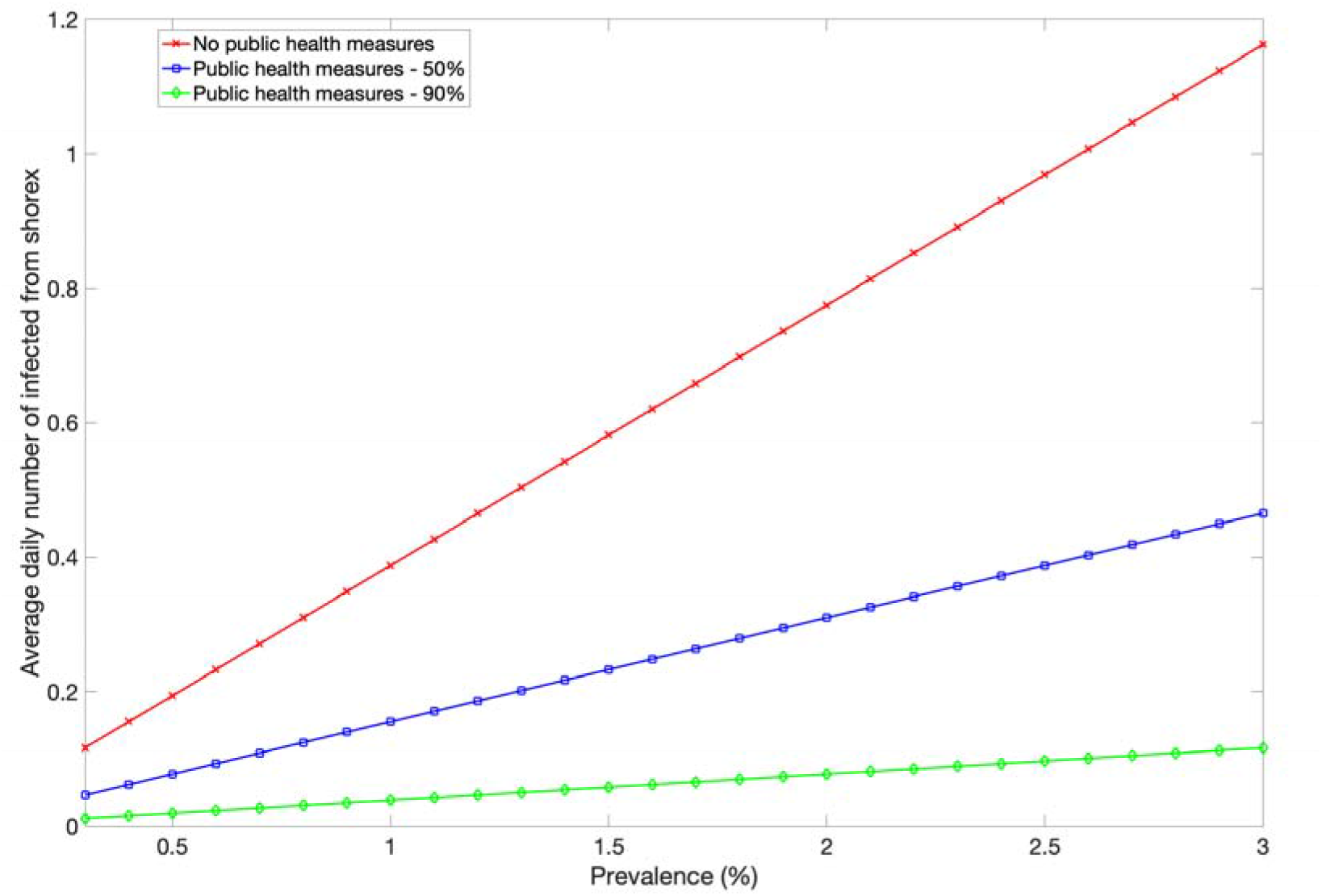
Illustration of the average daily number of infected individuals from shore excursions (shorex) as a function of the local prevalence and the extent of public health measures (e.g., facemask wearing). The number of infected passengers arising from daily shore expeditions depends on the number of susceptible passengers, the local reproduction number R0(shore), the local COVID-19 prevalence level, the amount of time that passengers spend on shore excursions (about 6 hours) as well as the extent of public health measures in place (e.g., facemask wearing). Here R0(shore) = 1.0. The extent of public health are modeled as a proportionate reduction in the transmission probability per contact which is assumed to vary from 50% to 90%. This is likely a conservative range that reflects the combined impact of enhanced surface sanitation, mandatory facemask wearing on board for passengers and crew, physical distancing protocols, air purifiers, and UV lights in all air handling units.

##### Global sensitivity and uncertainty analyses

To account for various model uncertainties including the epidemiology of the novel coronavirus, the characteristics of the cruises (duration, local prevalence) as well as the characteristics of the test on our modeling results (Table 1), we relied on global uncertainty and sensitivity analyses. Specifically, we performed global sensitivity and uncertainty analysis to quantify the effect of changes in the model parameters on the cumulative number of cases during the cruise. For sensitivity analyses, we rank model parameters according to the size of their effect on the total cases that occur during the duration of the cruise using partial rank correlation coefficients (PRCC)^39^. The larger the partial rank correlation coefficient, the larger the influence of the input parameter on the cumulative number of cases. All of the input parameters were sampled from uniform distributions (ranges given in Table 1) following Latin Hypercube Sampling with 200 samples.

**Table 1.** Parameter definitions, baseline values, and ranges considered in global sensitivity analyses

| Parameter definition | Baseline value | Range |
| --- | --- | --- |
| Number of crew | 465 (small ship) | 2,000 (big ship) |
| Number of passengers | 930 (small ship) | 4,000 (big ship) |
| Local prevalence | 1% | 0.3-3% |
| Mean peak timing for infectivity profile (days) | 6 | 5 - 7 |
| Length of the cruise (days) | 7 | 4-14 |
| Basic reproduction number aboard the cruise ship, $R_{0(ship)}$ | 12 | 9-16 |
| Basic reproduction number during shore excursions, $R_{0(shore)}$ | 1.0 | 0.6-1.6 |
| Extent of public health/social distancing measures (%) | 60% | 50-90% |
| Maximum PCR test sensitivity (%) | 85% | 80-95% |
| Time from infection to peak PCR test sensitivity (days) | 6 | 5-7 |

## Results

Figure 5 compares the mean and the 95% CI of the distribution of the caseload at the end of 7-day cruises for three different scenarios with and without interventions using the baseline parameter values shown in Table 1.

**Figure 5.**
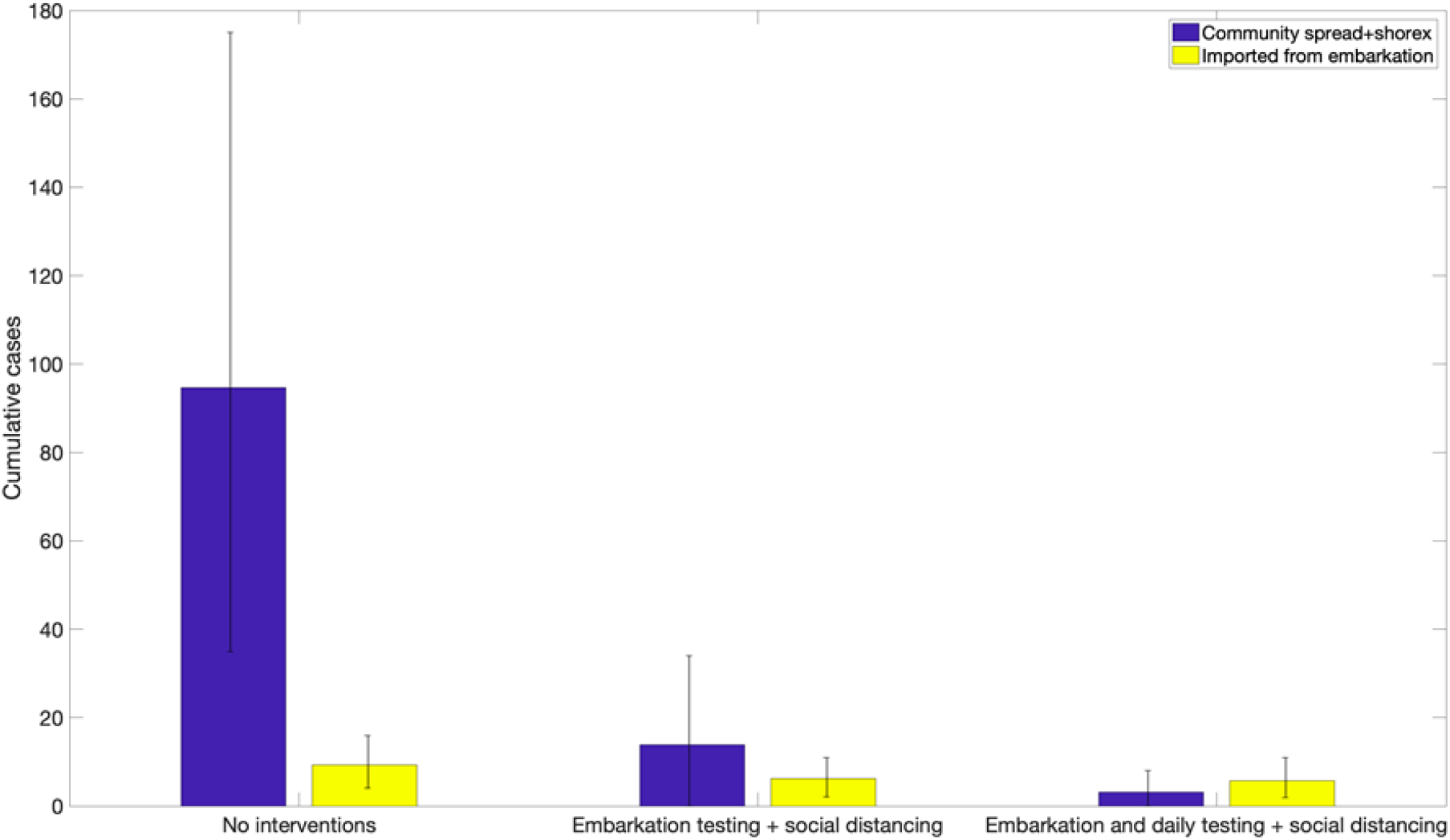
Distribution of the cumulative number of infections during 7-day cruises for three different scenarios using the baseline parameter values displayed in Table 1. The error bar reflect the 95% CI of the outcome distribution from 200 stochastic realizations.

- In the absence of interventions, the average number of infections during a 7-day cruise was estimated at 139.2 (95% CI: 43.8, 270) while the number of imported cases from embarkation was estimated at 9.6 (95% CI: 4.0, 15). This result underscores the substantial number of imported cases that lead to rapid growth in case numbers even when the baseline COVID-19 prevalence in the general population is 1 COVID-19 case in 100 people.
- In contrast, for the **embarkation testing** and public health measures scenario, we estimated an average of 14.9 cases (95% CI:1, 39.0) during the cruise whereas the number of imported cases from embarkation was estimated at 5.2 (95% CI: 1, 10). Hence, the average number of case importations is only reduced from 9.6 cases in the absence of interventions to an average of 5.2 cases. Indeed, embarkation testing may not be able to detect all of the infected individuals prior to boarding especially those in the early infection stages given our current understanding of the temporal sensitivity profile in the PCR test (Figure 3).
- For the scenario of **embarkation testing and daily testing of all individuals**, the number of cases during the cruise is reduced to an average of 2.9 cases (95% CI:0, 8). In fact, the stochastic simulation curves of the daily cumulative number of cases across the three scenarios indicate that the strategy that tests during embarkation together with daily testing of all individuals is the only strategy that rapidly flattens the growth trend in case numbers (Figure 6). This strategy greatly diminishes the probability that an infectious individual transmits the virus to other individuals aboard the vessel. While embarkation testing may not be able to detect all of the infected individuals prior to boarding, the great majority of infected individuals that are not detected during embarkation are likely to be identified soon after embarkation through daily testing strategies.

### Results from uncertainty and sensitivity analyses

#### In the absence of interventions

our results from the global uncertainty analyses after considering the uncertainty ranges for each parameter (Table 1) indicate that the average number of imported cases from embarkation is at 15 (95% CI:2, 32) while the average number of cases during the cruise including any cases during shore excursions (shorex) is 470 (95% CI:19, 1300). Furthermore, results from sensitivity analyses (Table 2 and Figure 7) indicate that the case load at the end of the cruise was most sensitive to the following parameters:

**Table 2.**
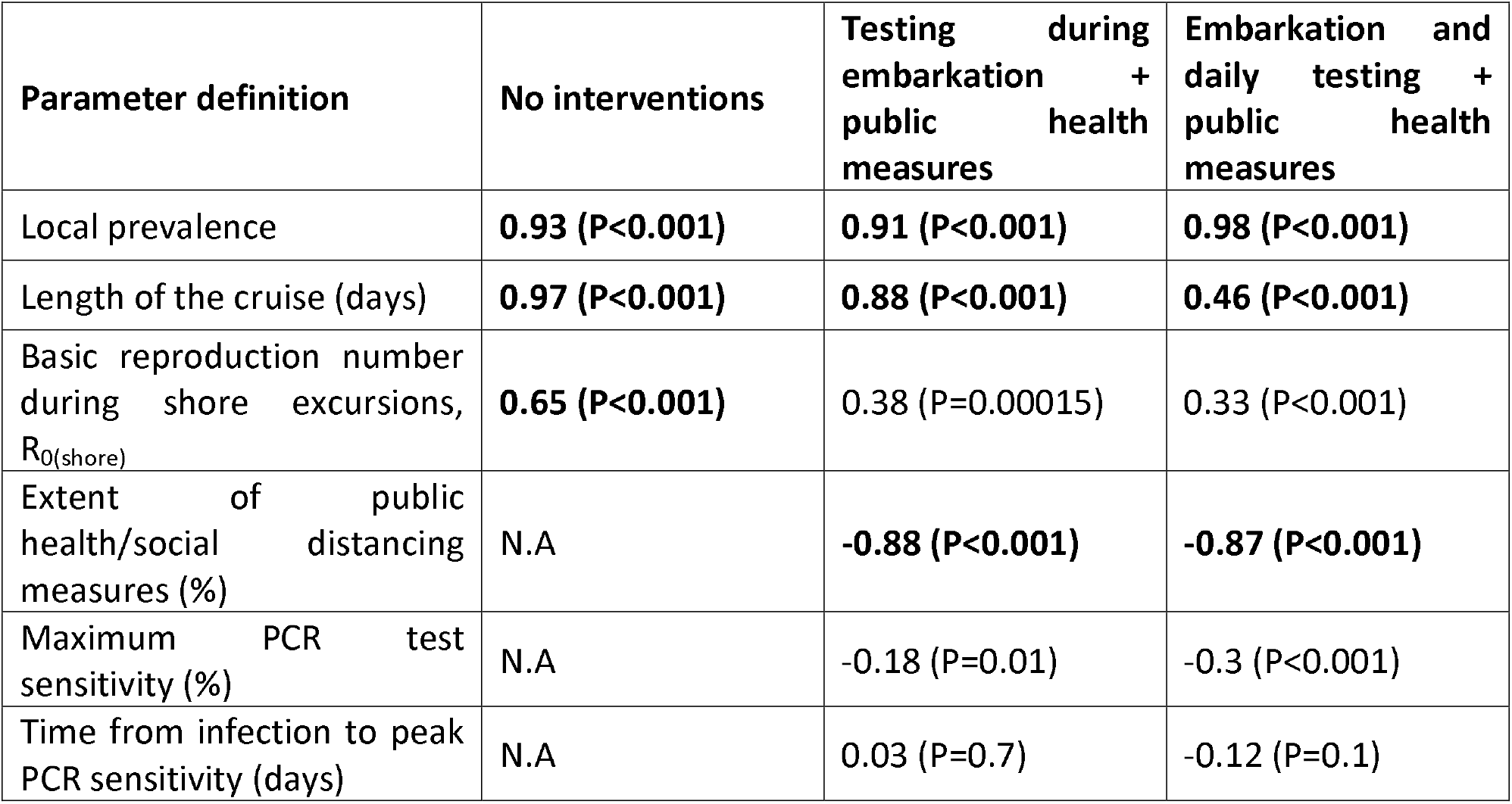
Partial Rank Correlation Coefficients (PRCC) derived from global sensitivity analyses that quantify the influence of the model parameters on the average final case load.

**Figure 6.**
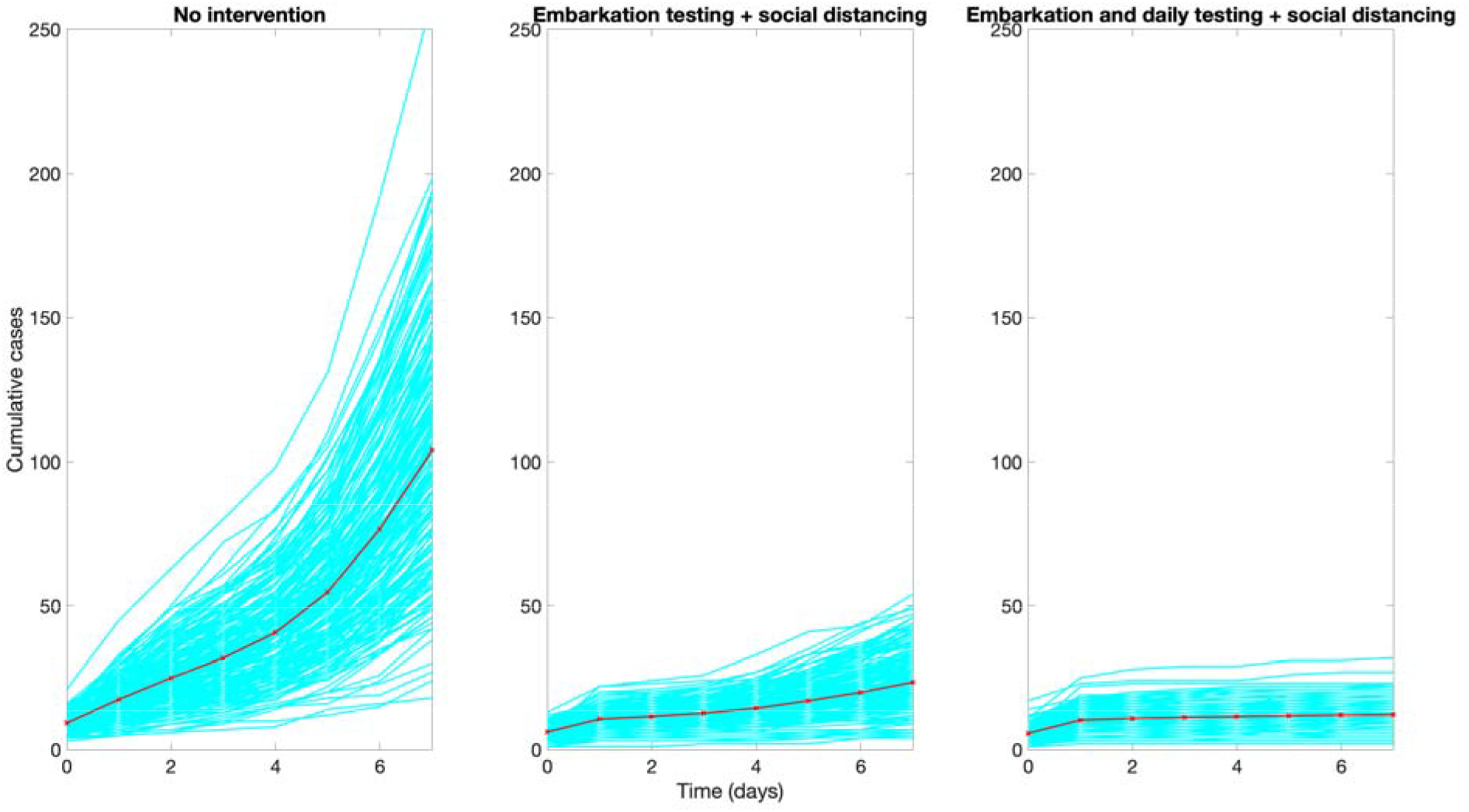
Epidemic curves of the cumulative number of infections during 7-day cruises for three different scenarios using the baseline parameter values displayed in Table 1. The red curve indicates the mean of the 200 stochastic realizations (cyan curves).

**Figure 7.**
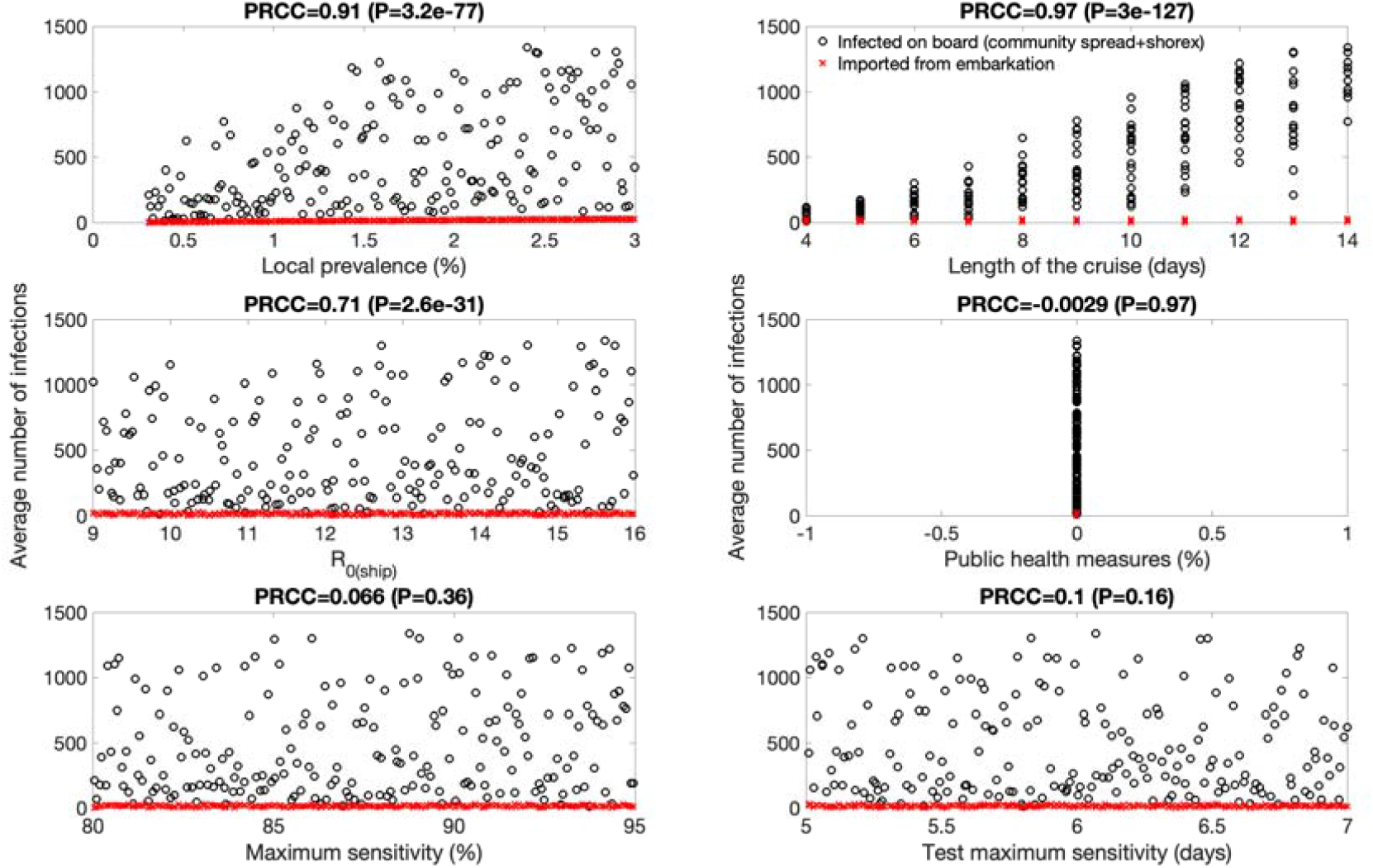
These scatter plots display the relationship between the model parameters and the average number of infections in the absence of testing strategies and public health measures. Partial rank correlation coefficients (PRCC) and their statistical significance (P value) after considering the uncertainty ranges for each parameter (Table 1) are also shown. Local prevalence and the length of the cruise are strongly and positively correlated with the expected number of cases at the end of the simulations. The basic reproduction number aboard the ship is also correlated with the average number of cases.

- The local community prevalence. This parameter is strongly correlated with the expected number of infected individuals boarding the vessel. Each infected individual that boards the vessel initiates a rapidly growing transmission chain given the high basic reproduction associated with the cruise environment.
- The duration of the cruise. This parameter directly modulates the time window during which susceptible may become exposed to infectious individuals on aboard the vessel. The basic reproduction number aboard the ship, R0(ship). This parameter is related to the likelihood that an infectious individual transmits the coronavirus to other individuals on aboard.

#### Embarkation testing and social distancing interventions

For the scenario that considers testing during embarkation together with public health measures, our results from the global uncertainty analyses indicate that the average number of imported cases from embarkation is at 9.6 (95% CI:1, 21) while the average number of cases during the cruise including any shorex cases is 45 (95% CI: 0, 260). Furthermore, results from sensitivity analyses (Table 2 and Figure 8) indicate that for this scenario the case load at the end of the cruise was most sensitive to the following parameters:

**Figure 8.**
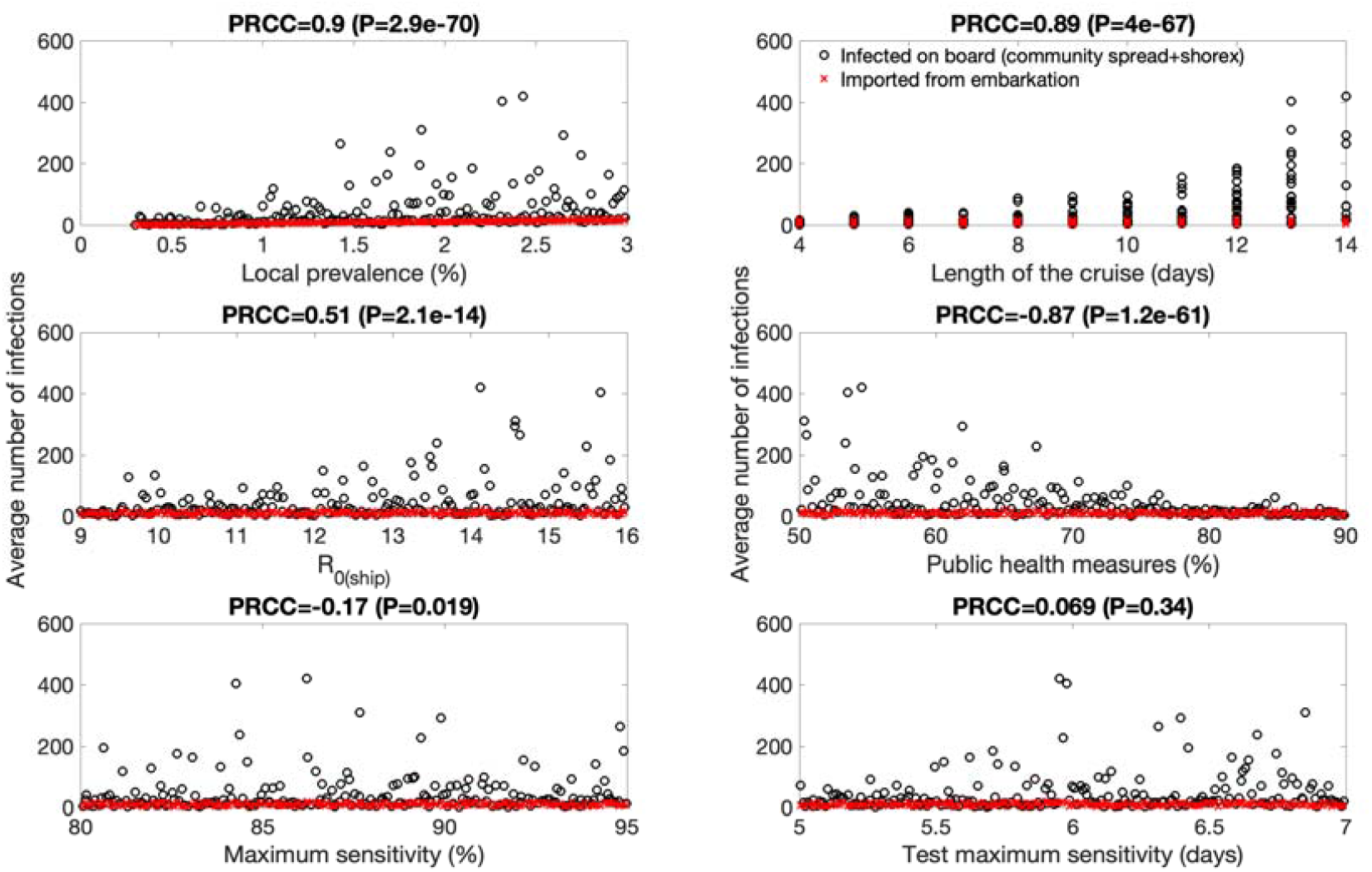
These scatter plots display the relationship between the model parameters and the average number of infections in the context of embarkation testing and public health measures. Partial rank correlation coefficients (PRCC) and their statistical significance (P value) after considering the uncertainty ranges for each parameter (Table 1) are also shown. Local prevalence, the extent of public health measures and the length of the cruise are highly and positively correlated with the expected number of cases at the end of the simulations. The basic reproduction number aboard the ship is only weakly correlated with the average number of cases.

- The local prevalence. Higher local prevalence levels increase the expected number of infected individuals boarding the vessel even when embarkation testing mitigates the number of infected individuals that make it aboard.
- The length of the cruise. Once an infected individual makes it aboard the vessel, the longer the duration of the cruise, the longer the time window during which susceptible individuals could be become exposed to infectious individuals aboard the vessel.
- The extent of public health measures. The transmission rate is mitigated proportionately by the level of public health measures, but it is unlikely to be reduced to zero.

#### Embarkation and daily testing + social distancing interventions

For the scenario that considers testing during embarkation and daily testing of all individuals together with public health measures, our results from the global uncertainty analyses indicate that the average number of imported cases from embarkation is at 9.6 (95% CI:1, 21) while the average number of cases during the cruise including any shorex cases is 4.7 (95% CI:0,16). Furthermore, results from sensitivity analyses (Table 2 and figure 9) indicate that for this scenario the case load at the end of the cruise was most sensitive to the following parameters:

**Figure 9.**
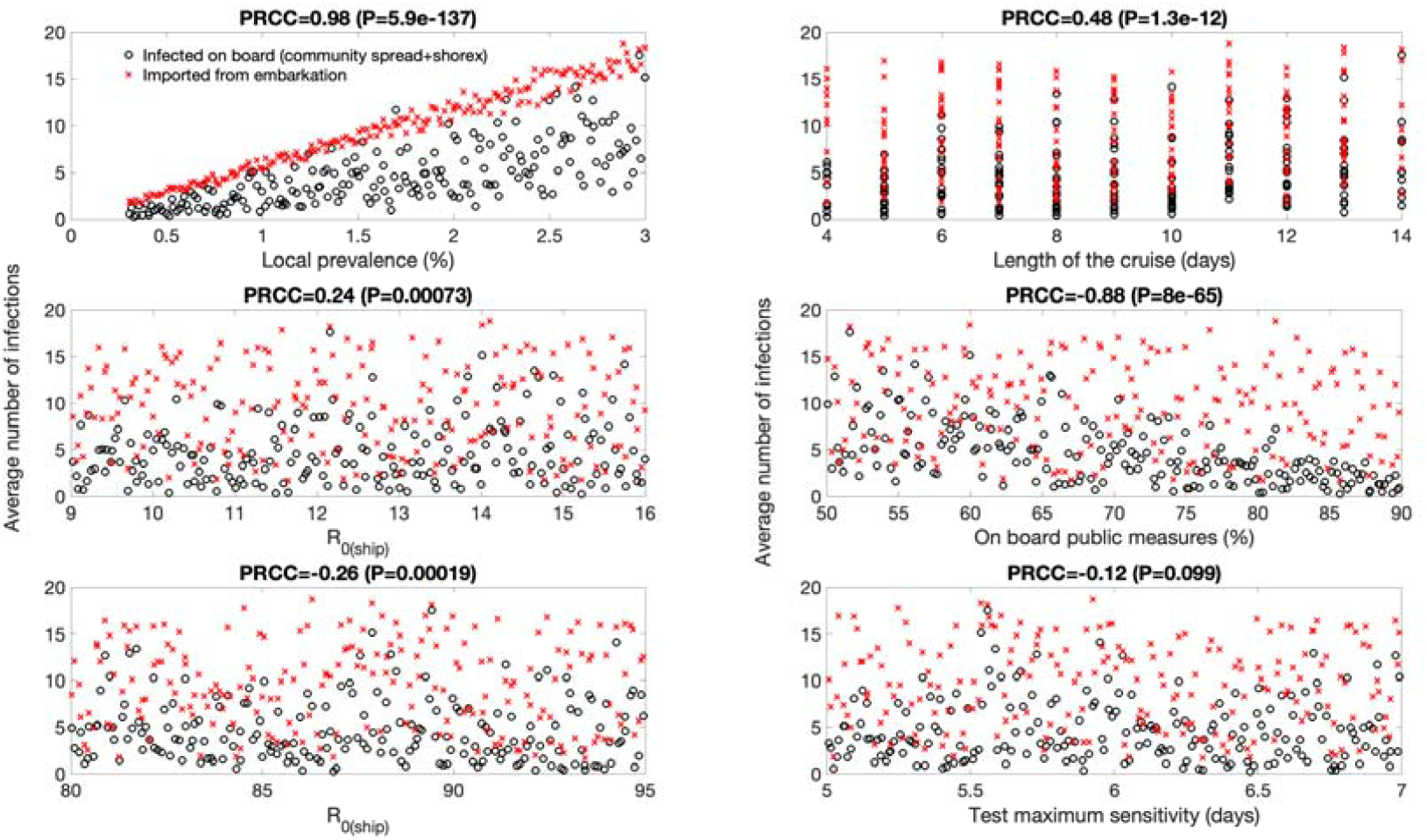
These scatter plots display the relationship between the model parameters and the average number of infections in the context of embarkation and daily testing together with social distancing protocols. Partial rank correlation coefficients (PRCC) and their statistical significance (P value) after considering the uncertainty ranges for each parameter (Table 1) are also shown. Local prevalence and the extent of public health measures are positively correlated with the expected number of cases at the end of the simulations. The basic reproduction number aboard the ship does not play a significant role while the length of the cruise is only weakly correlated and does not substantially affect the outcome.

- The local prevalence. Embarkation testing and daily testing greatly mitigate the average number of cases occurring on board the vessel compared to embarkation testing only (Figure 8). Yet, the number of cases during the cruise is still correlated with the local prevalence level.
- The extent of public health measures. The transmission rate during the cruise is negatively correlated with the extent of public health measures.

It is worth noting that compared to embarkation testing only, the length of the cruise does not play an important role in the average number cases expected during the cruise in this scenario.

## Discussion

Using an individual-level stochastic transmission model parameterized for COVID-19 transmission dynamics, we find that the implementation of PCR testing during embarkation and daily testing of all individuals aboard together with social distancing measures can greatly mitigate the number of cases during cruises for a wide range of parameter uncertainty related to the epidemiology of the novel coronavirus, PCR test characteristics relating to the sensitivity profile, and the conditions surrounding the operation of the cruises (local prevalence and cruise duration). Importantly, our findings also indicate that testing during embarkation is not sufficient to avert outbreaks even when moderate levels of social distancing measures are considered ^37^. Daily testing is key to minimize the number of infections irrespective of the duration of the cruise.

Cumulative incidence from the baseline model without considering the effects of interventions are significantly sensitive to variations in the local community prevalence (0.3-3%), the duration of the cruise ship (4-14 days), and the basic reproduction number aboard the cruise ship (9-16). For intervention scenarios incorporating testing strategies and social distancing, cumulative cases are most sensitive to the variation in local prevalence and the extent of social distancing measures (50-90%). Importantly, cumulative cases were not significantly sensitive to the variation assumed in the characteristics of the test sensitivity profile (Table 1).

The role of mathematical modeling has been instrumental in our current understanding of the transmission dynamics of COVID-19. Mathematical models have been employed in several previous studies to understand the effectiveness of non-pharmaceutical interventions to the control of SARS-CoV-2 in the Cruise ship setting such as Diamond Princess Cruise ship ^32,40-42^. While some of the previous studies have used the compartmental models that are limited in representing the full heterogeneity of contact patterns of people aboard ^32,40^, others that have employed the contact network models have not assessed the role of testing strategies in preventing COVID-19 outbreaks during the ocean cruise that involve shore excursions ^41,42^.

Our analysis is not exempt of limitations. First, there is still much uncertainty surrounding the full spectrum of infectiousness profiles in the population and additional data may prove useful to further refine this model component. Second, the contact network of all the individuals aboard the cruise ship was modeled following the small world network paradigm which provides a reasonable approximation to social interactions ^30^. Real-time collection of data on contacts occurring during cruises should help define a more accurate characterization of the contact dynamics that occur between crew members and passengers. Third, we model conservative scenarios since we do not account for potential effects arising from prior immunity shaped by past infections or vaccination campaigns. As COVID-19 vaccination campaigns are launched around the world, future studies could evaluate their impact on transmission dynamics.

## Data Availability

NA

## Acknowledgements

We thank Enrico Prunotto for helpful discussions on model development.

## Author contributions

GC analyzed the data and wrote the first draft of the manuscript. All authors contributed to model development, interpretation of the results and revising subsequent drafts of the manuscript.

## Figure legends

Figure 7. The timing of peak infectivity for each individual varies randomly, peaking at 5 (red curve),6 (blue curve), or 7 (green curve) days post infection according to a gamma distribution. We do not distinguish between symptomatic and asymptomatic individuals.

Figure 8. Contact network involving 1395 individuals on board the cruise ship following the small world network model with baseline values (K=8 and p=0.1). The dots represent individuals and the lines represent fixed contacts between individuals.

Figure 9. Illustration of the PCR test sensitivity profiles modeled according to a logistic function where a peak sensitivity of 80% is reached 5 (red curve), 6 (blue curve), or 7 (green curve) days post infection..

Figure 10. Illustration of the average daily number of infected individuals from shore excursions (shorex) as a function of the local prevalence and the extent of public health measures (e.g., facemask wearing). The number of infected passengers arising from daily shore expeditions depends on the number of susceptible passengers, the local reproduction number R0(shore), the local COVID-19 prevalence level, the amount of time that passengers spend on shore excursions (about 6 hours) as well as the extent of public health measures in place (e.g., facemask wearing). Here R0(shore) = 1.0. The extent of public health are modeled as a proportionate reduction in the transmission probability per contact which is assumed to vary from 50% to 90%. This is likely a conservative range that reflects the combined impact of enhanced surface sanitation, mandatory facemask wearing on board for passengers and crew, physical distancing protocols, air purifiers, and UV lights in all air handling units.

Figure 11. Distribution of the cumulative number of infections during 7-day cruises for three different scenarios using the baseline parameter values displayed in Table 1. The error bars reflect the 95% CI of the outcome distribution from 200 stochastic realizations.

Figure 12. Epidemic curves of the cumulative number of infections during 7-day cruises for three different scenarios using the baseline parameter values displayed in Table 1. The red curve indicates the mean of the 200 stochastic realizations (cyan curves).

